# Genomic epidemiology demonstrates spatially clustered, local transmission of *Plasmodium falciparum* in forest-going populations in southern Lao PDR

**DOI:** 10.1101/2024.07.15.24310466

**Authors:** Ying-An Chen, Eric Neubauer Vickers, Andres Aranda-Diaz, Maxwell Murphy, Inna Gerlovina, Francois Rerolle, Emily Dantzer, Bouasy Hongvanthong, Hsiao-Han Chang, Andrew A. Lover, Nicholas J. Hathaway, Adam Bennett, Bryan Greenhouse

## Abstract

While there has been significant progress in controlling falciparum malaria in the Lao People’s Democratic Republic (PDR), sporadic cases persist in southern provinces where the extent and patterns of transmission remain largely unknown. To assess parasite transmission in this area, 53 *Plasmodium falciparum* (Pf) positive cases detected through active test and treat campaigns from December 2017 to November 2018 were sequenced, targeting 204 highly polymorphic amplicons. Two R packages, *MOIRE* and *Dcifer*, were applied to assess the multiplicity of infections (MOI), effective MOI (eMOI), within-host parasite relatedness, and between-host parasite relatedness (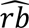). Genomic data were integrated with survey data to characterize the temporal and spatial structures of identified clusters. The positive cases were mainly captured during the focal test and treat campaign conducted in 2018, and in the Pathoomphone area, which had the highest test positivity and forest activity. About 30% of the cases were polyclonal infections, with over half of theses (63%) showing within-host relatedness greater than 0.6, suggesting that cotransmission rather than superinfection was primarily responsible for maintaining polyclonality. A large majority of cases (81%) were infected by parasites genetically linked to one or more other cases. We identified five genetically distinct clusters in forest fringe villages within the Pathoomphone district, characterized by a high degree of genetic relatedness between parasites (mean 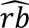 = 0.8). Four smaller clusters of 2-3 cases linked Moonlapamok and Pathoomphone districts, with an average 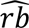 of 0.6, suggesting cross-district transmission. Most of the clustered cases occurred within 20 km and 2 months of each other, consistent with focal transmission. Transmission clusters identified in this study confirm the role of ongoing focal parasite transmission occurring within the forest or forest-fringe in the highly mobile population.

**Author summary:** In low-transmission settings, integrating genomic data could help capture fine-scale dynamics in circulating parasites that may not be easily detected by traditional surveillance. Our work demonstrates that even with very few malaria cases, incorporating active surveillance and genomic analysis can help track residual transmission pockets and routes, including among highly mobile populations whose infection sources are historically difficult to trace. Transmission clusters identified in this study provided clear evidence of ongoing focal transmission of *Plasmodium falciparum* in southern Laos. The addition of high-resolution genomic data to active surveillance can aid in targeting and assessing interventions in similar areas approaching elimination.

## Introduction

The Lao PDR has made remarkable progress in its efforts to eliminate malaria, with a substantial decrease in malaria cases from an estimated 280,000 in 2000 to approximately 2,300 cases in 2022, representing a 99% decline [1, 2]. As the country has approached elimination, malaria transmission has become more heterogeneous and exhibited greater geographic and socio-demographic clustering [3-5]. In northern Lao PDR, malaria transmission is very low, with a predominance of *Plasmodium vivax* infections. In contrast, southern Lao PDR bears over 90% of the nationwide malaria disease burden with co-endemic transmission of both *Plasmodium falciparum* and *Plasmodium vivax* [6, 7].

The higher malaria burden observed in southern Lao PDR can be attributed to several factors. These include increased exposure to malaria vectors in remote or forested areas, limited access to public health services, and the difficulty in identifying and improving interventions for high-risk populations, particularly mobile forest workers [8, 9]. As a result, the Center for Malariology, Parasitology in Laos have focused their prevention efforts on forest-goers and remote villages, with the aim of accelerating *P. falciparum* elimination by 2024 [10]. Although *P. falciparum* infections in Laos have declined due to active interventions, residual transmission persists in southern Laos [11, 12] and little is known regarding specific transmission pockets and routes.

In this study, we employed a genomic epidemiology approach to investigate the transmission patterns of *P. falciparum* cases identified during an active screening trial in southern Lao PDR. This method allowed us to explore the genetic diversity of infecting parasites, assess the relatedness among the identified infections, and identify the likely spatial and temporal extent of transmission, providing valuable insights into the dynamics of malaria transmission in these remaining high-risk areas.

## Methods

### Ethics statement

Ethical approval was obtained from the Ministry of Health review board in Lao PDR (reference numbers 2016-014 and 2017-075), and from the UCSF IRB (reference numbers 16-19649 and 17-22577). Informed written consent was obtained from all participants who were interviewed and had dried blood spots (DBS) collected. For participants under the age of 18, consent was provided by their caregivers, and children aged 10 and above also provided their own assent directly.

### Study design

Samples and data were obtained from active malaria test and treat campaigns conducted in four administrative districts, Pathoomphone (PT), Moonlapamok (MP), Sanasomboon (SB), and Sukhuma (SK) in Champasak province from 2017 to 2018 [13, 14]. The campaigns included a baseline survey conducted between November and December 2017, a village-based mass test and treat (MTAT) campaign conducted in June to July 2018, and an endline survey conducted in October to November 2018. A continuous focal test and treat (FTAT), enrolling a few forest goers daily, was conducted between the baseline and endline surveys, from March to November 2018 [14]. A total of 5749, 2904, 18144, and 7870 individuals were enrolled in the baseline, FTAT, MTAT, and endline surveys, respectively (S1 Table).

The survey data included basic characteristics including age, gender, diagnostic results, date of collection, residential district and village, history of fever, taking of anti-malarial drugs, and forest activities. During the trial, rapid diagnostic tests (RDTs) were used for active screening of parasite infected cases [13]. A total of 65 samples were confirmed as *P. falciparum* infections (S1 Table), and their corresponding dried blood spots (DBS) were sent to UCSF for storage and subsequent DNA extraction, qPCR, and amplicon sequencing.

### Laboratory methods

Malaria DBS were DNA extracted using Tween-Chelex method described in Teyssier et al., 2021 [15]. Briefly, a 6-mm disc of the DBS punch was washed overnight in 0.5% Tween 20/1X PBS solution at 4°C. A second wash was performed using 1X PBS for 30 min at 4°C. After removing PBS, the DBS punch was covered with 10% Chelex 100 resin in water and incubated for 10 min at 95°C, then centrifuged at 15,000 rpm for 10 min. The resulting supernatant, containing the DNA, was transferred into a PCR tube and stored at -20°C.

Parasite density was measured using the *varATS* qPCR protocol designed by Hofmann et al [16]. A series of DNA standards, extracted from mock DBS with known concentrations of 1, 10, 100, 1,000, and 10,000 parasites per microliter, was utilized for qPCR standardization. Duplicate sets of the standard series were employed to generate a standard curve for each run, with a linearity (R^2^) exceeding 0.98 considered as a validated run. The parasite density of the samples was determined by interpolation using the constructed standard curve.

Sensitive amplification of 204 targets was performed with modifications to the CleanPlex protocol by Paragon Genomics, CA, USA [17]. The genotyping panel was designed to include microhaplotype targets with high diversity in Southeast Asian parasites in an analogous manner to our original panel targeting parasites in sub-Saharan Africa [18] (details available in the Appendix file 2, Paragon panel ID PGD375). Following amplification, the samples were subjected to bead purification, quantification, and subsequent pooling based on parasite density. The pool was then sequenced with 150 bp paired-end clusters on an Illumina sequencer. We performed three replicates of library construction for each sample, refining conditions in an effort to optimize amplification efficiency, enhance sequence recovery, and ensure reproducibility.

### Bioinformatic pipeline and quality check of sequences

The targeted amplicon data were processed using a pipeline v0.0.8 that involved Nextflow [19] and DADA2 [20] (available at https://github.com/EPPIcenter/mad4hatter). This pipeline consisted of core modules encompassing adapter removal, quality control, sequence inference using DADA2, and post-processing, optionally involving the masking of low complexity regions.

Sequenced data from three batches were merged, and for each detected allele, the one with the most reads was selected. The output allele data was then used for downstream analysis. From our sequenced data, we successfully recovered a total of 180 distinct amplicons from the 200-plex oligo pools after filtering out amplicons with no reads. To ensure data quality, we implemented two criteria: (1) the total reads for each sample should exceed the number of amplified amplicons multiplied by 30, and (2) a minimum locus coverage (where reads per locus were non-zero) of 50 out of 180 (S1 Fig). Sequenced data that passed both the quality check criteria were retained for downstream genetic measurements.

### Data analysis

After retrieving the filtered post-pipeline polyallelic data, we utilized the *MOIRE v3.1.0* R package to implement a Markov chain Monte Carlo (MCMC)-based approach for estimating the multiplicity of infection (MOI) [21]. Here, MOI represents the number of genetically distinct parasite strains within an individual, accounting for genotyping error and within-host relatedness. Two additional metrics were also estimated by *MOIRE*, the within-host parasite relatedness (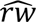) and the effective MOI (eMOI) [21]. The effective MOI (eMOI) adjusts the MOI for underlying within-host relatedness (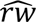), which is the average proportion of the genome that is identical by descent across all strains within the same host. The eMOI is defined as (MOI – 1) * (1 – 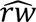) + 1 [21]. Evaluating eMOI and within-host relatedness is particularly relevant in low transmission settings as it helps detect signals of inbreeding within the parasite population and allows for more accurate estimates of MOI. In this study, polyclonal infections were defined for infections estimated to have an eMOI greater than 1.1. Expected heterozygosity and allele frequencies were also estimated using *MOIRE*.

To estimate between-host relatedness, we applied the *Dcifer v1.2.0* R package, a method to calculate pairwise genetic distance as identity by descent (IBD) between infections, including those which are polyclonal [22]. By inputting the MOI and allele frequency estimated from *MOIRE*, each pair obtained an estimation of relatedness ranging from 0 to 1 and a corresponding statistical significance (p-value). Genomic clusters were then identified by applying a relatedness cut-off of 0.3 or greater, ensuring that all captured pairs had a significance level at 0.05 (S2 Table and S2 Fig). The composition and connectivity of each genomic cluster were visualized using the *igraph* v1.3.5 R package [23].

Genomic data were integrated with survey data to characterize the temporal and spatial structure of identified clusters. Spatial mapping of identified genomic clusters was visualized using *QGIS* v3.22.7, and the spatial distance between cases was calculated by the Haversine formula, the shortest distance between two points on a sphere using their latitudes and longitudes. Pairwise time differences were calculated based on the interval in days between the sampling dates of each pair. Univariate logistic regression analysis was used to identify risk factors contributing to sample clustering. The factors included in the analysis were: age, gender (with female as the reference), parasite density group (with < 100 parasites/uL as the reference group), mixed infections with P. vivax, district (districts outside Pathoomphone as the reference), having fever, use of anti-malarial drugs, and staying overnight in the forest.

## Results

### Within-host and population diversity in southern Lao PDR

Sixty-five positive cases were identified, with most captured during the FTAT campaign and in the Pathoomphone area. Of these, 53 (82%) successfully passed the sequence quality check and were retained for downstream analysis (S1 Table). About 60% of the infected individuals were asymptomatic (non-feverish), and over 90% of the cases were male adults aged between 15 and 74, with a median age of 27 years (Table 1).

**Table 1.** Characteristics of all, clustered and non-clustered positive cases. In addition to the age variable, numbers in the second to forth columns represent sample counts, and numbers in brackets indicate the proportions in each group. † denotes variables with missing values. The variable “feverish” has 2 missing values, and the variable “took antimalarials” has 9 missing values in the overall data. * Unadjusted p-values are estimated by univariate logistic regression analysis. Significant p-values (< 0.05) are in *Italic*. Two significant factors, collection year and staying overnight in the forest, were further tested with permutation test, and only individuals who spent overnight in the forest remained significant (p = 0.005) to the clustering membership.

| Characteristic | All | Clustered | Non-clustered | Unadjusted p-value* |
| --- | --- | --- | --- | --- |
| **No.** | 53 | 43 | 10 |  |
| **Age median [Q1-Q3]** | 27 [22-38] | 27 [22-37] | 34.5 [21-42] | 0.73 |
| **Gender** |  |  |  |  |
| Male (%) | 48 (91) | 39 (91) | 9 (90) | 0.95 |
| Female (%) | 5 (9) | 4 (9) | 1 (10) |  |
| **Parasite density** |  |  |  |  |
| < 100 p/uL (%) | 17 (32) | 12 (28) | 5 (50) |  |
| ≥ 100 p/uL (%) | 36 (68) | 31 (72) | 5 (50) | 0.19 |
| **Mixed infection with *P. vivax*** | 9 (17) | 7 (16) | 2 (20) | 0.78 |
| **Year** |  |  |  |  |
| 2017 (Baseline, %) | 9 (17) | 5 (12) | 4 (40) |  |
| 2018 (MTAT & FTAT, %) | 44 (83) | 38 (88) | 6 (60) | 0.04 |
| **District** |  |  |  |  |
| Pathoomphone (%) | 44 (83) | 37 (86) | 7 (70) | 0.24 |
| Moonlapamok (%) | 7 (13) | 6 (14) | 1 (10) |  |
| Sanasomboon (%) | 1 (2) | 0 (0) | 1 (10) |  |
| Sukhuma (%) | 1 (2) | 0 (0) | 1 (10) |  |
| **No. of unique villages** | 21 | 12 | 9 |  |
| **Feverish (%)†** | 21 (40) | 19 (44) | 2 (20) | 0.22 |
| **Took antimalarials (%)†** | 11 (21) | 11 (21) | 0 (0) | 0.99 |
| **Stayed overnight away from home village in the forest (%)** | 46 (87) | 40 (93) | 6 (60) | 0.01 |

The genetic diversity across 180 loci in this isolated setting with low transmission had an average expected heterozygosity (He) of 0.3, though 36 loci still had moderate to high diversity with H_E_ > 0.5 (S3 Fig). Within-host genetic diversity was low; in most infections a minority of loci had more than one allele detected (S1B Fig). Consistent with these data, the majority (70%) of cases were estimated to be monoclonal infections with the remaining polyclonal infections having a maximum MOI of 3 (S4 Fig). The population mean MOI was estimated to be 1.6 (95% CI = 1.3 - 1.9, Fig 1A).

**Fig 1.**
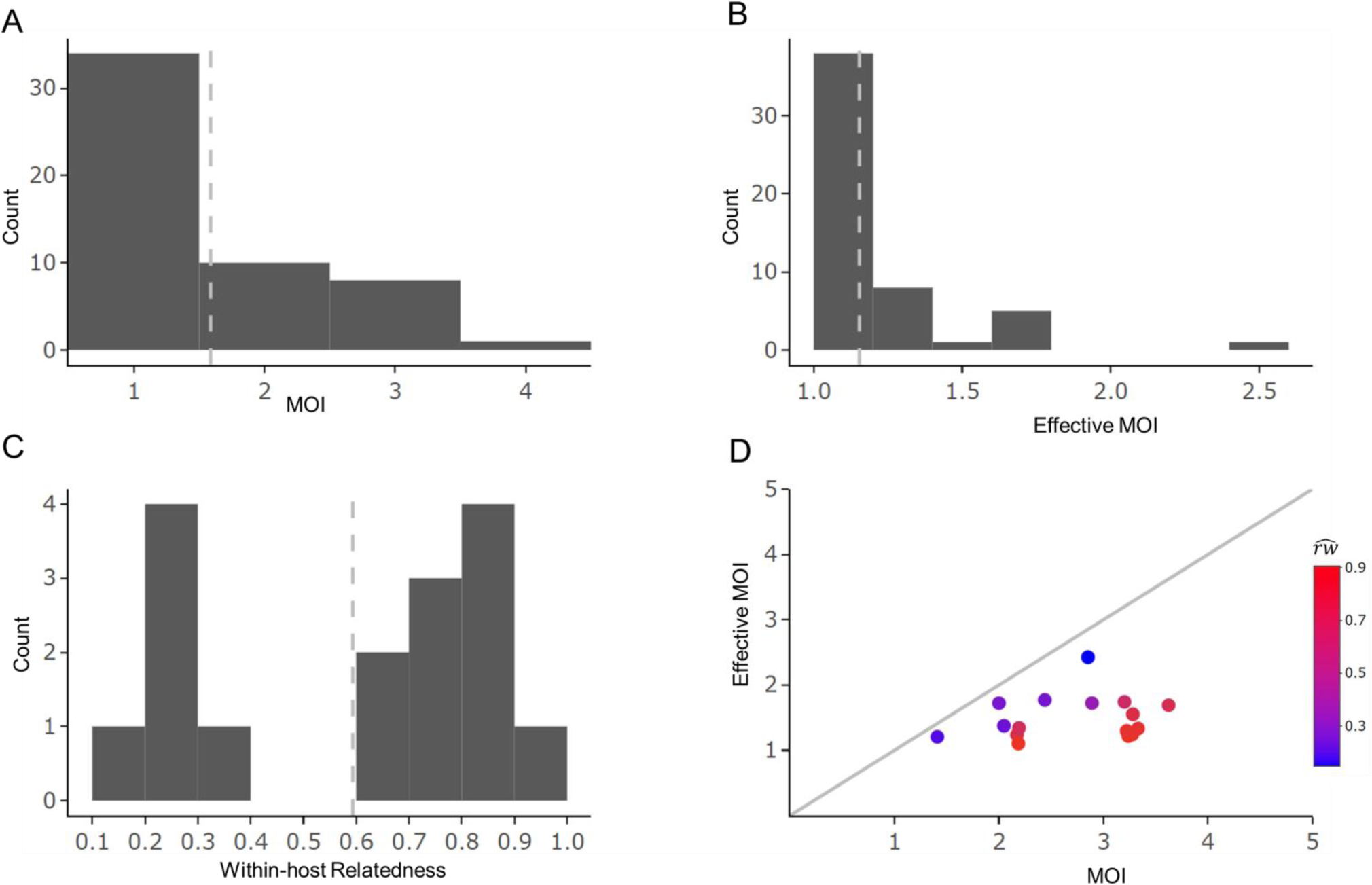
Distribution of mean posterior estimates of MOI, effective MOI, and within-host relatedness. The mean of posterior estimates for each sample were obtained using MOIRE. The dashed gray lines in plots A, B and C represent the population mean. Plots C and D show the estimates of within-host relatedness (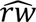) for polyclonal samples (16/53, 30% of total cases), and the relationship with MOI and effective MOI estimates.

The identified 16 polyclonal infections exhibited an average within-host relatedness of 0.6, with a bimodal distribution (Fig 1C). Ten of the 16 polyclonal infections showed within-host relatedness above 0.6 (range = 0.64 - 0.91), while the remaining 6 had within-host relatedness less than 0.4 (range = 0.15 - 0.37). This indicates that it was common for distinct parasite strains within hosts with polyclonal infections to share a common ancestry, likely due to serial co-transmission and inbreeding of strains. The extent of within host relatedness reduced the population MOI to an average effective MOI (eMOI) of only 1.15 (Figs 1B and 1D).

### Genomic clusters identified in the forest-going population

A total of 53 samples formed 1,378 distinct inter-sample pairs. Among these, 125 pairs (9.1%) showed a relatedness greater than 0.3; all sample pairs above this level of relatedness had statistical support for being genetically related more than expected by chance (p < 0.05). Using a relatedness cutoff of 0.3, 43 (81%) cases had infections related to at least one other infection. The fact that most cases were related to others suggests ongoing local transmission in the identified cases. These genetically linked infections formed five main clusters (labelled A to E), as well as four smaller clusters (labelled F to I, Fig 2). The factor associated with infections being related to others was whether an individual had stayed overnight away from their home village in the forest (Table 1, unadjusted OR = 8.9, 95% CI = 1.6 - 49.9, p = 0.01). Due to collinearity between the collection year and the FTAT survey that enrolled the most forest goers, we performed a permutation test on both the collection year and overnight forest-visiting status to assess their relationship to clustering membership. After 1000 permutations, spending overnight in the forest remained significant (p = 0.005), while equal subsampling of the 2017 and 2018 data repeatedly showed no significant relationship to clustering membership (p = 0.66). This suggests that overnight forest activity was associated with a higher risk of being locally infected and/or transmitting parasite clones within the recent past.

**Fig 2.**
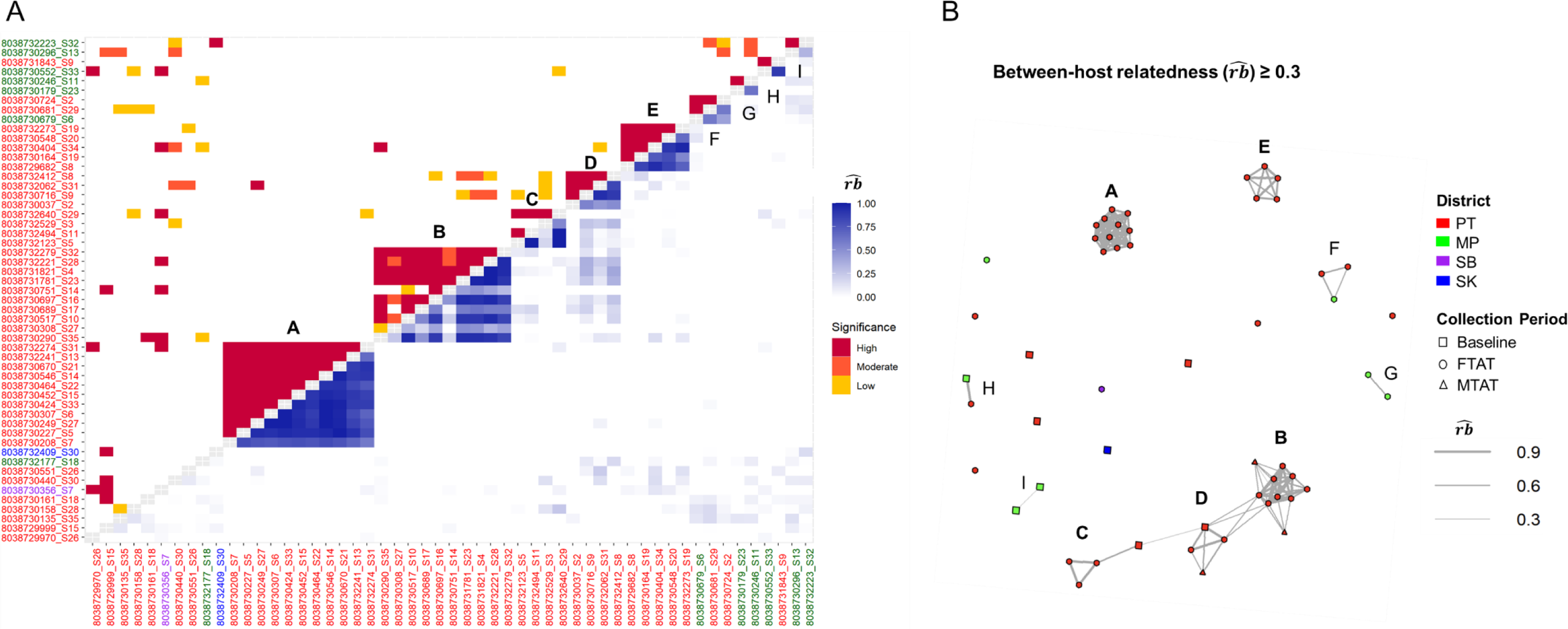
Between-host relatedness estimated by *Dcifer*. (A) Estimates from the lower triangle represent between-host relatedness (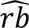) across all parasites in each pair of infections, where 0 signifies completely unrelated infections, and 1 signifies completely related infections. The upper triangle indicates significance level: high significance (p < 0.001), moderate significance (0.001 ≤ p < 0.01), low significance (0.01 ≤ p < 0.05). Cases are ordered by clustered levels, and the colors of labels indicate the districts of origin for positive cases (PT = Pathoomphone, MP = Moonlapamok, SB = Sanasomboon, and SK = Sukhuma). Five large, highly related clusters (A to E marked in bold) with high statistical significance are identified, along with four smaller clusters (F to I). (B) Network plot: Lines connect nodes (cases) representing pairs with significant relatedness above 0.3, and line width reflects the strength of relatedness. Node colors correspond to the districts of positive cases, also indicated by the axis labels in plot A. Different shapes represent sample collection periods: Baseline (2017/11-12), Focal Test and Treat (FTAT, 2018/03-11), and Mass Test and Treat (MTAT, 2018/06-07).

Related cases showed strong spatial and temporal clustering (Figs 3 and 4). For large and highly related clusters A through E, the spatial distances between clustered cases were relatively short (cluster means ranging from 5 to 11 km) while the smaller, less related cross-district clusters F, H, and I had larger distances. Most clusters also had a relatively short average time difference between sample pairs of 2 months.

**Fig 3.**
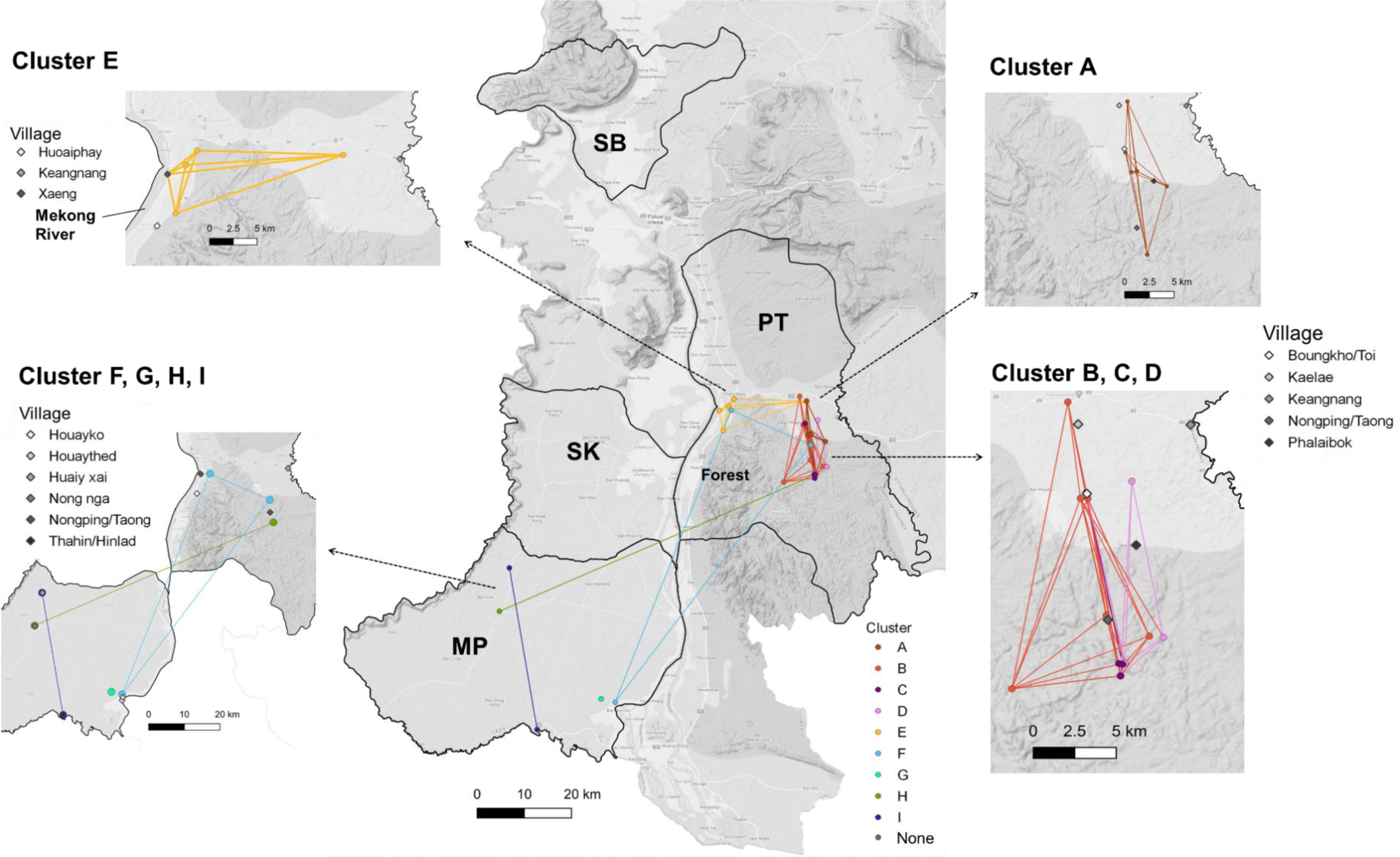
Spatial distribution of genomic clusters. Malaria cases were identified in four districts: Pathoomphone (PT), Moonlapamok (MP), Sanasomboon (SB), and Sukhuma (SK). The dots on the map represent the locations where clustered cases were collected, while the diamonds indicate the residential villages from which they originated. Clusters A, B, C, and D are concentrated in the eastern region of the Pathoomphone district, near the fringes of the forest. Cluster E is situated in the western part of the Pathoomphone district, close to the Mekong River. Four smaller clusters F, G, H, and I were identified between the Moonlapamok and Pathoomphone.

**Fig 4.**
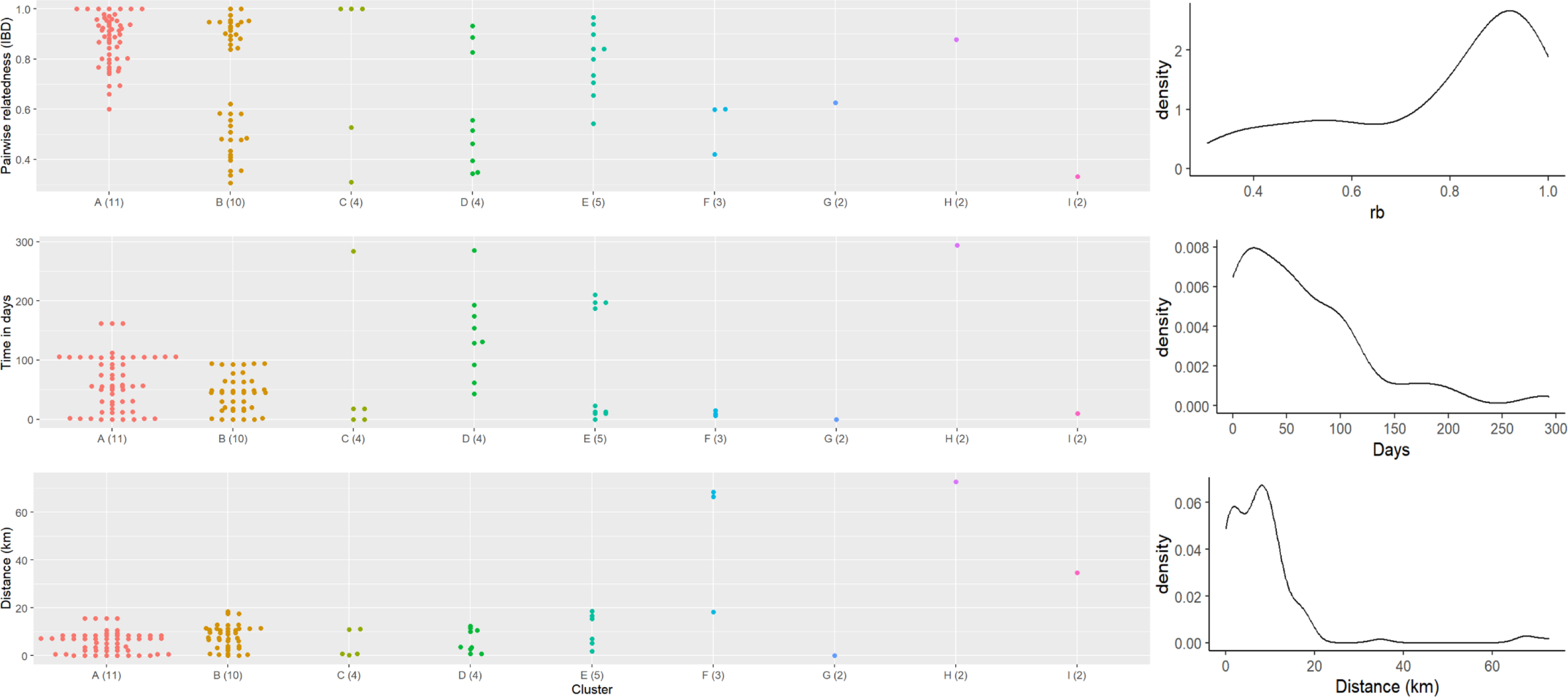
Pairwise relatedness, time difference, and spatial distance within each cluster. Genomic clusters were identified using a pairwise relatedness cutoff of 0.3. The x-axis represents each cluster, and the sample size is indicated in parentheses. Day interval is computed as the difference between the sampling dates of two positive cases. Spatial distance is calculated using the Haversine formula, the shortest distance between two points on a sphere using their latitudes and longitudes. The overall distribution of all three variables is shown on the right.

Large and highly related clusters showed different spatial distributions, with clusters A to D primarily located on the east side of the Pathoomphone district, while cluster E was mainly distributed in the western part of the Pathoomphone district, near the Mekong River (Fig 3). Cluster A comprised 11 cases centered at the Phalaibok village and showed high relatedness (mean 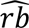 = 0.87). Clusters B, C, and D were comprised of 18 cases and were predominantly found in the Bougkho/Toi and Nongping/Taong villages. These three clusters were weakly interconnected by 1 to 2 samples from a more distant time point, with a relatedness of 0.3. Thus, when the relatedness cutoff was increased to 0.4 or more, they were identified as individual clusters (S5 Fig). Cluster E primarily involved individuals from Xaeng village who had visited the forest and were identified in April 2018, but with only one individual residing on the east side of the Pathoomphone district and identified 6 months later than the other cases (mean 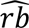 = 0.8). These results are compatible with some cases within a cluster being related to each other by direct transmission (highly related) and others indirectly linked from common ancestors and few intermediate unobserved infections (less highly related).

Clusters F to I, each consisting of 2-3 cases, were identified between the districts of Pathoomphone and Moonlapamok. Clusters F and H showed connections across district boundaries, while clusters G and I were both located within the Moonlapamok district. These clusters displayed a lower degree of relatedness compared to the clusters found in the Pathoomphone district, with an average pairwise relatedness of 0.58. Except for cluster H, all pairs within these clusters had a short time span of 0-15 days. Although a longer temporal gap was detected in cluster H (294 days), their relatedness remained high (0.88) in this pair, indicating sustained transmission possibly through unobserved intermediate infections between the Moonlapamok and Pathoomphone districts over the course of a year.

## Discussion

This study offers valuable insights into the transmission patterns of falciparum malaria in Southern Laos. The within-host diversity of the *P. falciparum* population in this region was found to be limited, with infections primarily characterized by monoclonal infections or highly inbred polyclonal infections consistent with sustained low-level local transmission. Most cases, particularly those with a history of overnight stays in the forest, had parasites closely related to several others in the area and could be categorized into one of several distinct genetic clusters. These data provide further evidence of sustained focal transmission and highlight geographic areas of potential transmission. The identified connections consistent with local transmission were mainly distributed along the forest fringes, emphasizing the risk of malaria transmission in those who reside near the forest or frequently visit it.

The clusters of transmission identified in this study were consistent with previous findings indicating that the at-risk group primarily consisted of male adults who spent the night in the forest, with their activities centered around villages located near the forest [14]. Adding precision to this general finding, we noted that the majority of potential transmission links were temporally related and spatially concentrated within a 20-kilometer radius on both the east and west sides of the Pathoomphone district, an area previously noted to have high levels of forested and agricultural activities [7]. In addition, we also detected a few cross-district transmission links between the Pathoomphone and Moonlapamok districts, which may be attributed to mobility patterns observed among the forest-goer population. One plausible explanation is sustained transmission within the extensive forested area in Pathoomphone district, seeding other areas via primary roadways connecting both districts. This forested area in Pathoomphone serves as a hub for local and potentially more distant individuals engaged in forest-related livelihood activities such as hunting. We also found some longer time spans (4 to 10 months) within these cross-district clusters, suggesting the existence of undetected intermediate reservoirs, potentially asymptomatic individuals who contributed to sustained transmission but were not captured in our active surveillance campaigns.

Our study highlights the value of integrating parasite genomic data into malaria surveillance for uncovering transmission patterns, identifying transmission hotspots, and revealing previously unrecognized transmission routes [24, 25]. *P. falciparum* cases were sporadic in our study region and were detected with active surveillance, but while details of individuals’ behaviors were collected, evidence regarding how and where infections were acquired and the geographic and temporal scales of transmission were mostly unknown. In our study, we employed a genomic epidemiological approach made possible by a series of recently developed technologies, including multiplexed amplicon sequencing of diverse microhaplotypes [18, 26] and methods for analysis of parasite diversity and relatedness which appropriately account for the biology of Pf including sexual recombination and polyclonal infections [21, 22]. Amplicon sequencing allows for deep coverage of highly polymorphic genomic regions, enabling the detection of minor clones and more accurate characterization of polyclonal infections [18, 24, 27]. In our study, this technique enabled the simultaneous recovery of abundant reads from approximately 200 amplicons in DBS with a wide range of parasite density, including one-third with a parasite density of < 100 parasites/μL. Unbalanced reads and systematic bias in estimated genetic metrics due to parasite density were not prevalent in our results (S6 Fig). Analysis methods accommodated polyalleleic data along with probabilistic measures of uncertainty and provided statistical inference, allowing us to conduct a principled exploration of spatial and temporal transmission patterns by characterizing the genetic relatedness of cases.

In summary, our study successfully showcases a genomic epidemiological strategy that augmented active surveillance with genomic data to reinvestigate the transmission dynamics of falciparum malaria in Champasak province in Southern Lao PDR. This was achieved despite having a small number of positive falciparum infections. The identified routes and foci of residual *P. falciparum* transmission can offer valuable insights for targeted interventions in the study area.

## Data Availability

All data produced in the present work are contained in the manuscript

https://drive.google.com/drive/folders/1e15s3Ns2BXZc_ZJlJmDkGkoU2vEHiNp1?usp=sharing

## Acknowledgements

We would like to thank the study participants and study teams for their cooperation, and the funding support by the Bill and Melinda Gates Foundation [OPP1116450] to MEI/UCSF.

## Author contributions

YAC designed the study, conducted the analysis, and wrote the manuscript. AB, HHC, and BG supervised the analysis. ENV and AAD performed the experiments and collected the laboratory data. FR, ED, BH, AAL, and AB designed the parent study and led data collection. NJH and AAD contributed to the amplicon sequencing panel. MM and IG developed the analysis packages.

## Supporting information

### Appendix file 1

S1 Fig. Quality control (QC) of sequenced positive samples.

S2 Fig. Distribution of 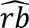 p-value. with (A) the corresponding p-value and (B) the log10-transformed

S3 Fig. Expected heterozygosity in 180 highly polymorphic loci.

S4 Fig. Median MOI and eMOI in each sample.

S5 Fig. Clustering network with different between-host relatedness cutoffs.

S1 Table. Number of positive and sequenced samples.

S2 Table. Pairwise relatedness and proportion of significant pairs.

### Appendix file 2

Amplicon information for Paragon panel ID PGD375.

### Appendix file 3

Meta-data of analyzed 53 Pf-positive samples, including survey data, sequenced data, and *MOIRE* results.

### Appendix file 4

Drug resistance analysis of 53 Pf-positive samples

