## Appendix file 1: Supplementary figures and tables for "Genomic epidemiology demonstrates spatially clustered, local transmission of *Plasmodium falciparum* in forest-going populations in southern Lao PDR"

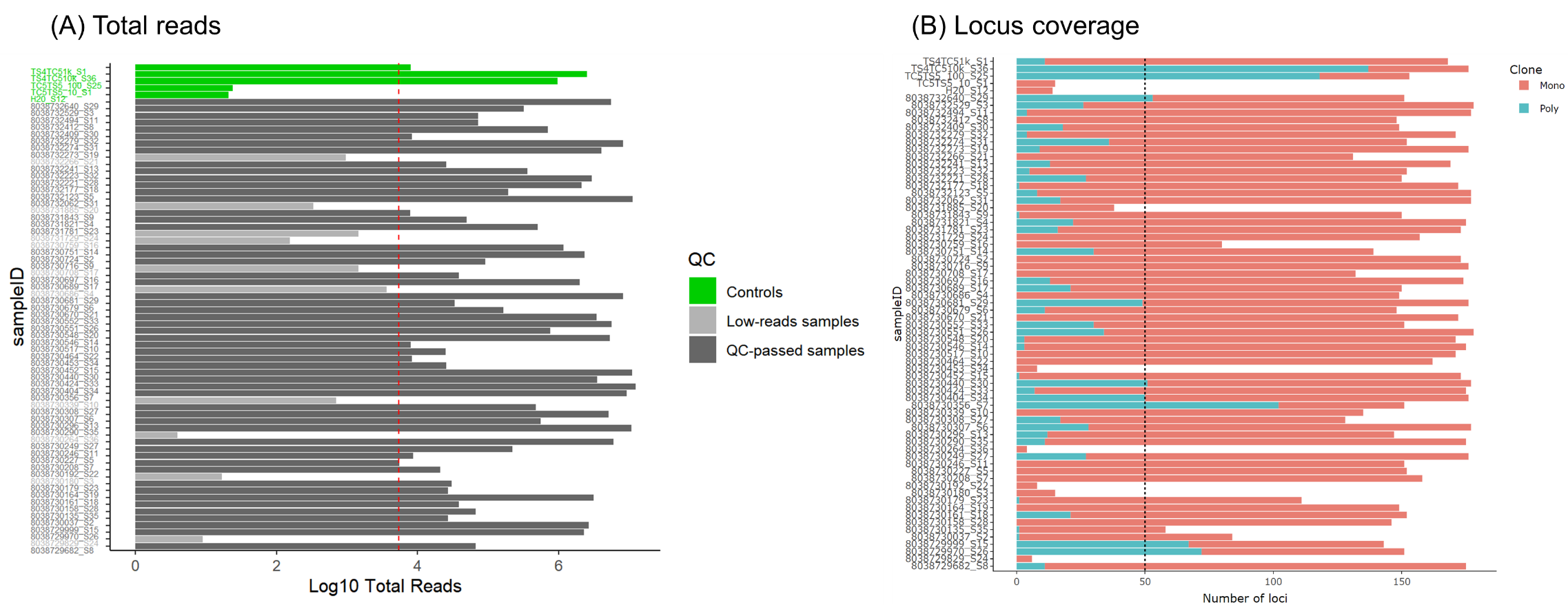

**S1 Fig. Quality control (QC) of sequenced positive samples.** A total of 65 positive cases were subjected to sequencing and recovering 180 diverse loci. Out of these, 53 samples (82%) successfully passed the quality control using two filters: (A) Total read counts > total number of loci * 30, (B) Locus coverage > 50 out of 180 (Note: "mono" denotes a locus with a single allele, while "poly" indicates a locus with more than one allele). The dashed lines in both plots represent the filtering thresholds. Only the samples that passed both thresholds were retained for downstream analysis.

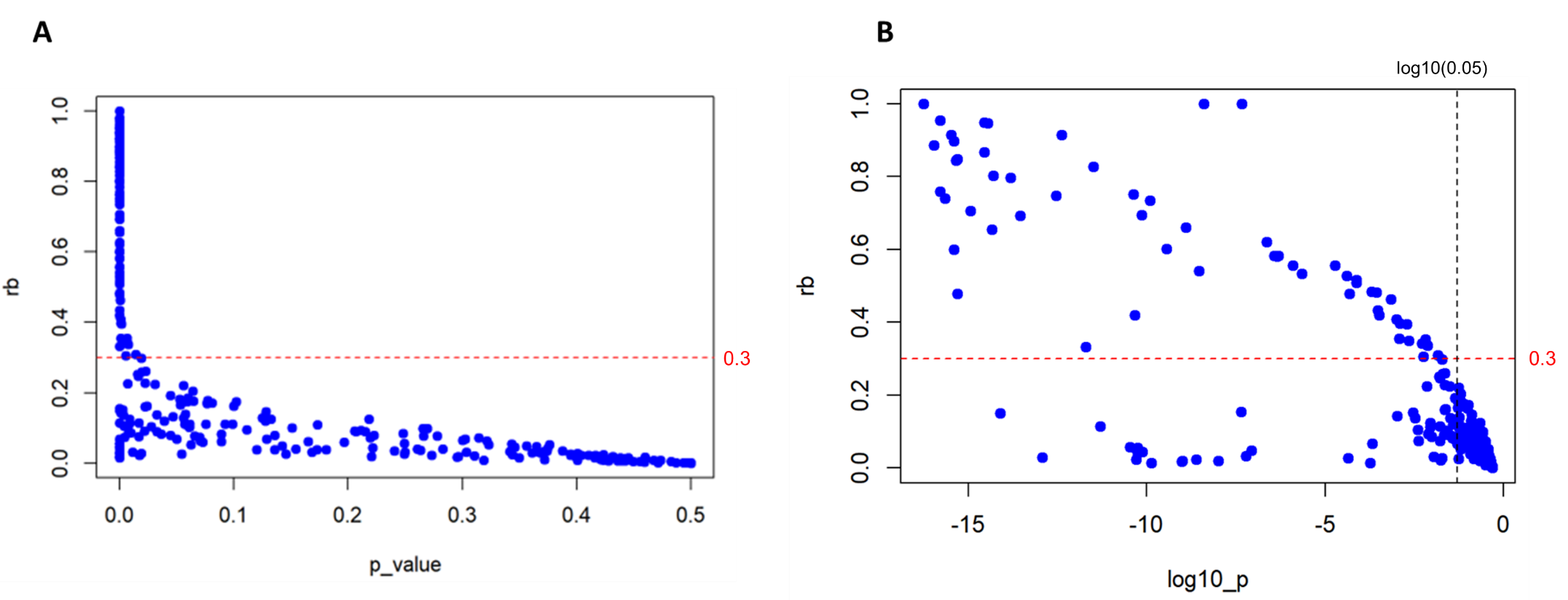

**S2 Fig.** **Distribution of** $\hat{\boldsymbol{rb}}$ **with (A) corresponding p-value and (B) log10-transformed p-value.**

The p-values are divided by two for one-sided tests. The variation of the p-values for $\hat{rb}$ ≤ 0.3 (red dashed line) became larger, resulting in more non-significant pairs. Therefore, a minimum $\hat{rb}$cutoff of 0.3 (red dashed line) was used to define related pairs with a significance level of 0.05 (black dashed line), where all the pairs with $\hat{rb}$ ≥ 0.3 fit.

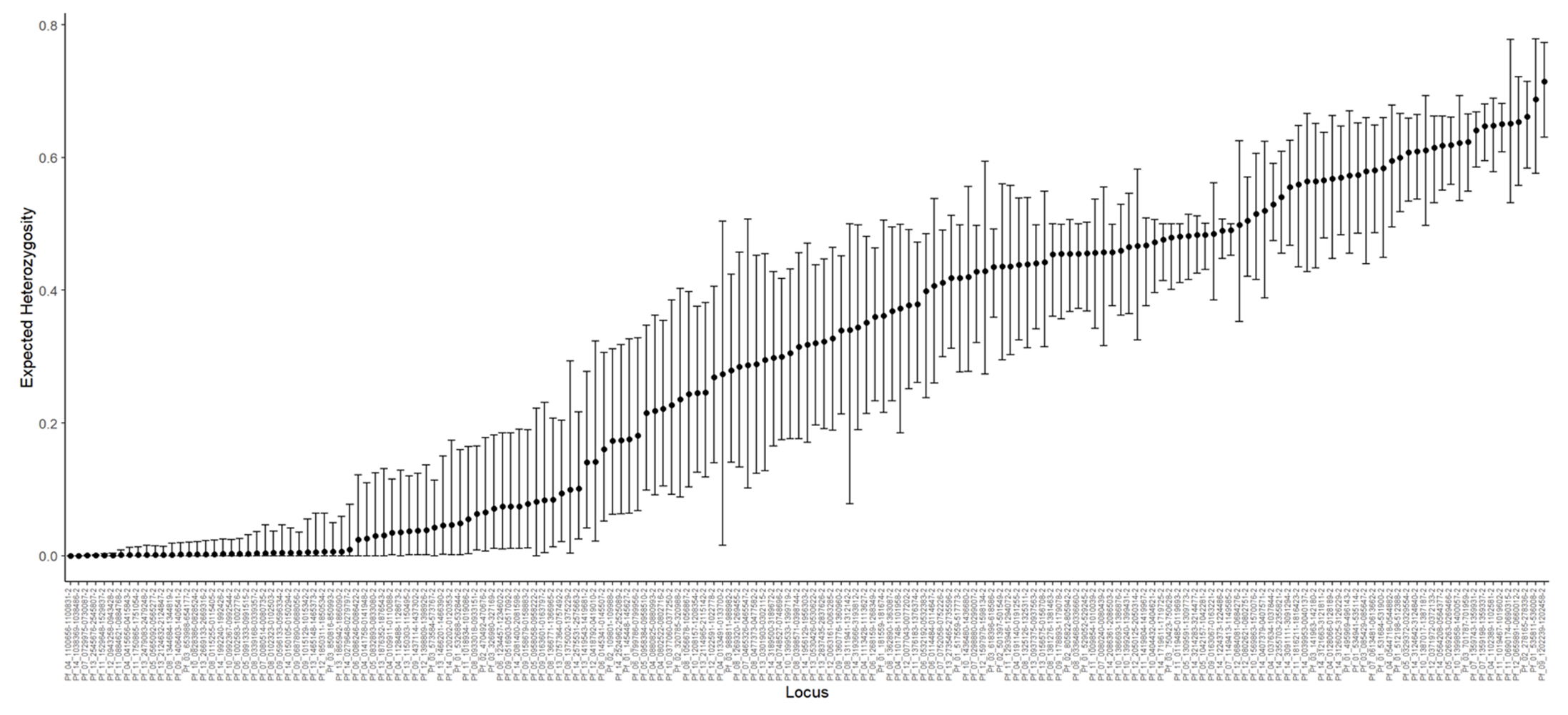

**S3 Fig. Expected heterozygosity in 180 highly polymorphic loci.** The dots indicate the mean estimates, and the bars show 95% credible interval.

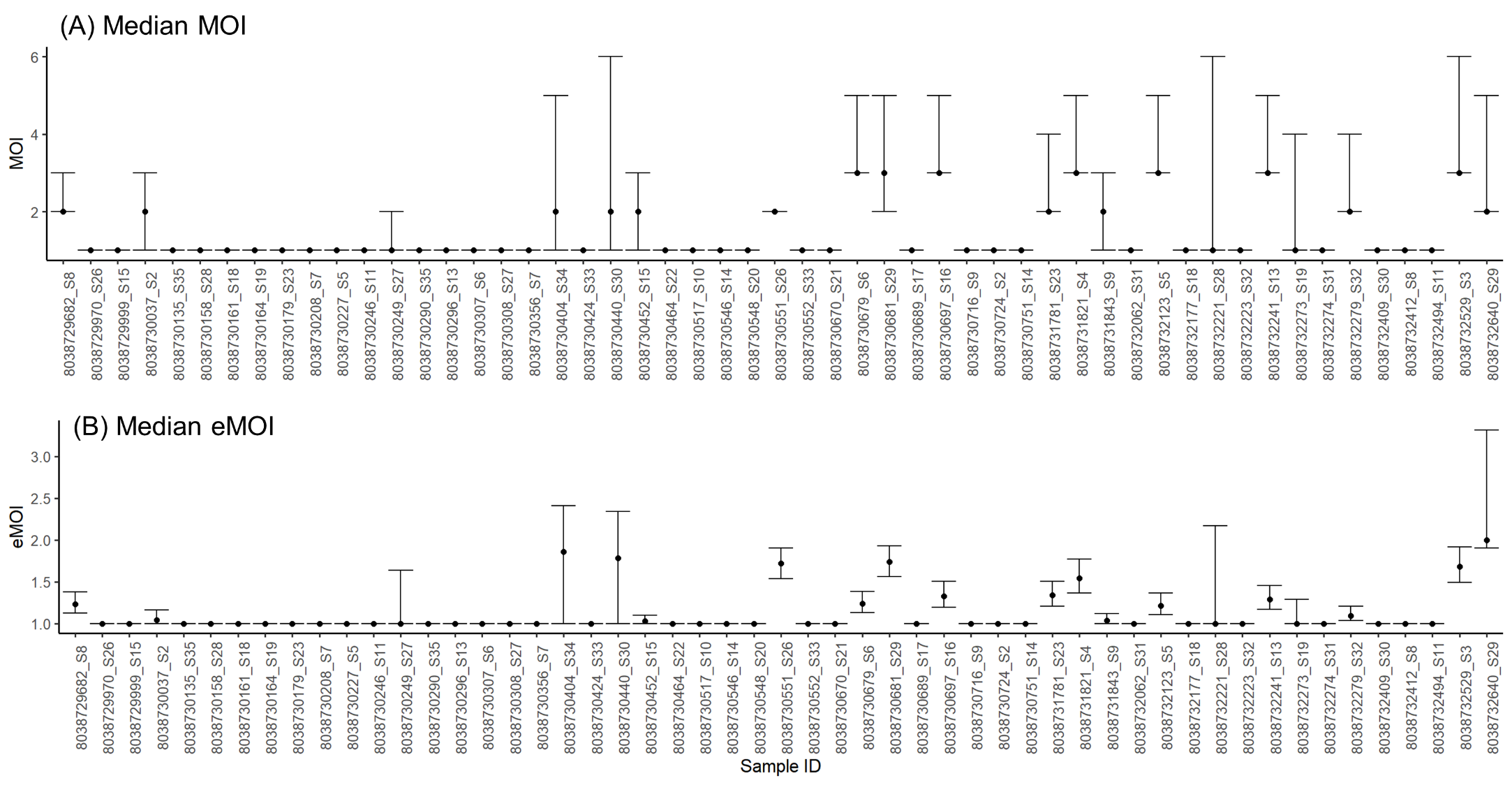

**S4 Fig. Median MOI and eMOI in each sample.** The upper plot displays the median MOI, while the lower plot represents the median effective MOI. The bars show 95% credible interval of the posterior distribution of MOI and eMOI, and the dots indicate the median.

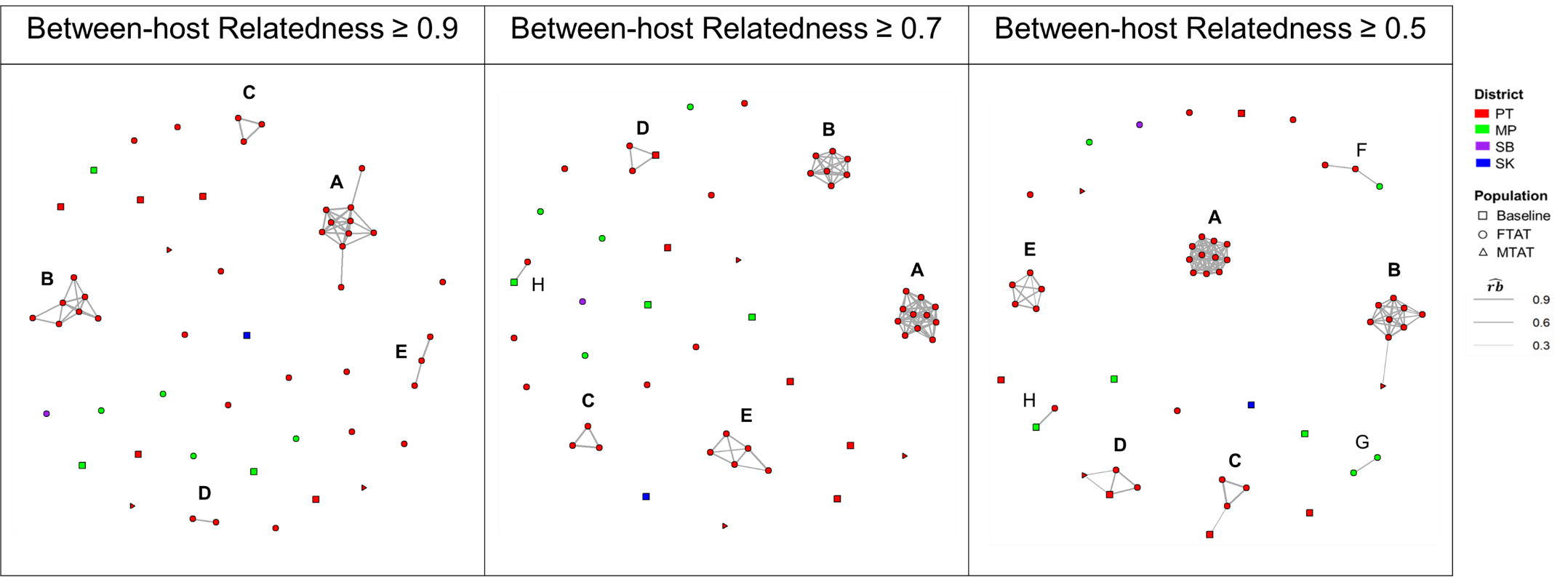

**S5 Fig. Clustering network with different between-host relatedness cutoffs.**

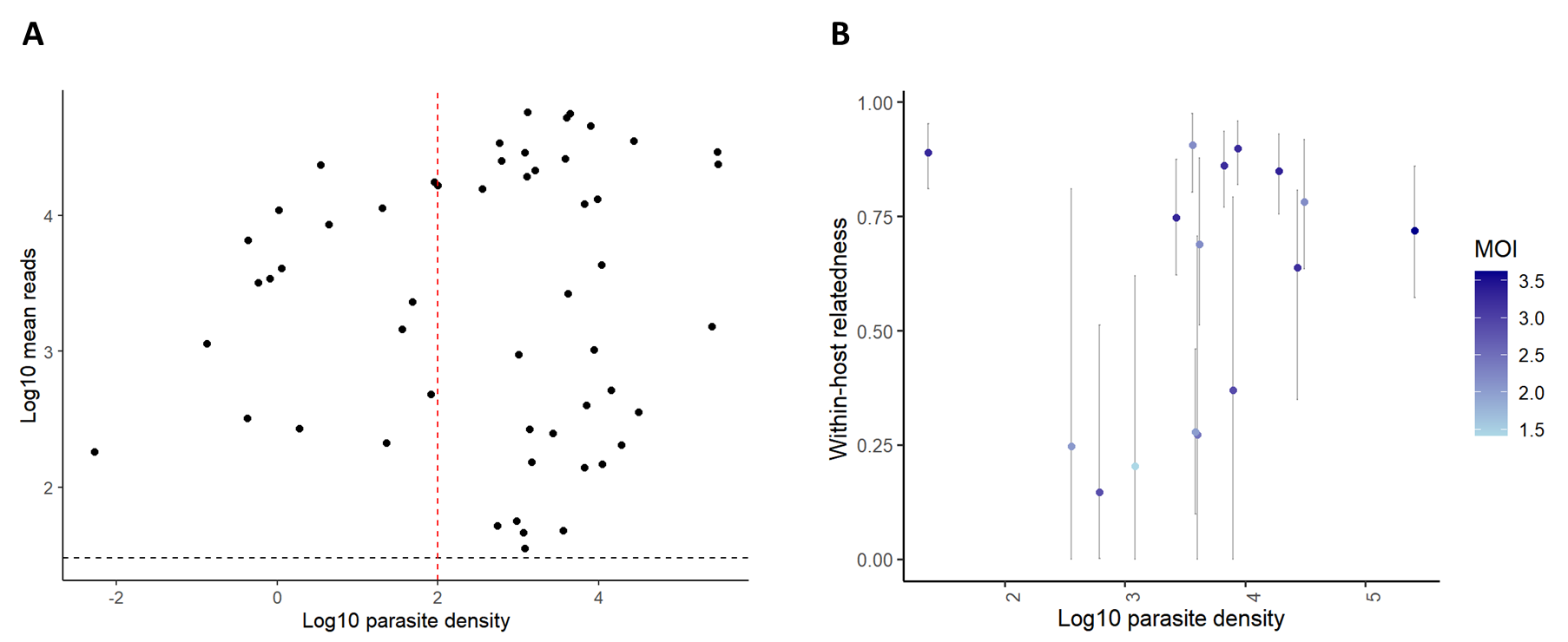

**S6 Fig. Relationship between parasite density, sequenced reads (A), and within-host relatedness (B).**

Parasite density was measured using the *varATS* qPCR. (A) In low-density samples with <100 parasites/uL (red dashed line), read counts performed similarly to high-density samples, with an average of at least 30 reads per amplicon (black dashed line), passing our quality control criterion. (B) Parasite density and within-host relatedness showed no significant correlation (Pearson’s p-value = 0.37). However, the confidence interval for within-host relatedness can be larger when the estimated within-host relatedness is lower and/or MOI is lower. This suggests that the uncertainty of within-host relatedness can be influenced by MOI estimates, rather than parasite density.

**S1 Table. Number of positive and sequenced samples.**

| **Survey (Time)** | **No. of collection** | **No. of PCR Pf-positive samples** | **No. of usable sequenced samples** |
| --- | --- | --- | --- |
| Baseline (Nov. to Dec. 2017) | 5,749 | 14 | 9 |
| FTAT (Mar. to Nov. 2018) | 2,904 | 48 | 41 |
| MTAT (Jun. to Jul. 2018) | 18,144 | 3 | 3 |
| Endline (Oct. to Nov. 2018) | 7,870 | 0 | 0 |
| **Total (%)** | **34,667** | **65 (0.19%)** | **53 (0.15%)** |

**S2 Table. Pairwise relatedness and proportion of significant pairs.**

| **Relatedness** | **Significant pairs  (p < 0.05)** | **Total pairs** | **% of significantly related pairs** |
| --- | --- | --- | --- |
| 0.0 – 0.1 | 27 | 1197 | 2 |
| 0.1 – 0.2 | 17 | 45 | 38 |
| 0.2 – 0.3 | 9 | 11 | 82 |
| 0.3 – 0.4 | 10 | 10 | 100 |
| 0.4 – 0.5 | 9 | 9 | 100 |
| 0.5 – 0.6 | 12 | 12 | 100 |
| 0.6 – 0.7 | 7 | 7 | 100 |
| 0.7 – 0.8 | 12 | 12 | 100 |
| 0.8 – 0.9 | 28 | 28 | 100 |
| 0.9 – 1.0 | 47 | 47 | 100 |

There is a total of 1,378 (53^2^ – 53 / 2) distinct sample pairs. We applied a significance level at p < 0.05.
